# Personalized Prediction of Regional Brain Atrophy in Parkinson’s Disease through Longitudinal AI Modeling

**DOI:** 10.64898/2025.12.29.25343190

**Authors:** Yusen Wu, Tatjana Rundek, Phuong Nguyen, Yelena Yesha, Matthew Feldman, Priyanka Atit, Jacob Miller, Tara Najafi, Ihtsham Haq

## Abstract

Parkinson’s disease (PD) involves variable patterns of brain atrophy in different motor and cognitive regions that differ across patients in both location and progression rate. Existing clinical tools and single-modal imaging methods can be difficult to analyze how individual patient’s atrophy will progress. Thus, we developed and evaluated deep learning models capable of understanding the pattern of 216 features of PD patients and predicting future regional brain atrophy in individuals with PD using longitudinal 3D MRI and clinical features. Our models combining long short-term memory (LSTM) networks and deep learning classification models to capture time-dependent changes in 29 PD-related brain volumes. Our model demonstrated accurate prediction of regional brain atrophy. In the classification models, 11 regions achieved AUROC greater than 80%. Regression results showed that our model produces an average MAE of 0.4395 and multiple regions *R*^2^ are more than 0.8. In conclusion, PD-related regional brain atrophy can be forecast with high test accuracy in longitudinal research cohort. These proof-of-concept results support the feasibility of developing personalized prognostic tools that integrate longitudinal 3D imaging with clinical data.

## 1 Introduction

Predicting the progression of Parkinson’s disease (PD) remains a critical challenge in clinical neurology due to the disease’s heterogeneous nature. Rates and patterns of brain atrophy vary widely among PD patients but align with clinical worsening [1]: Cortical thinning and subcortical volume loss are associated with more rapid motor and cognitive decline and ventricular expansion increases with advancing disease.

Personalized prediction of region-specific atrophy may offer a quantitative marker of future clinical progression. Traditional prediction tools have relied on patient clinical scales in isolation or a single imaging modality [2, 3, 4, 5] and often fail to capture individualized patterns of neurodegeneration. Recent advances in machine learning and artificial intelligence (AI) offer promising solutions by integrating clinical and radiological data from multiple time points into a unified predictive model [6]. This allows the model to learn patterns of all clinical features and predict future PD severity scores.

Current PD AI models have focused on classifying PD severity [7, 8, 9, 10, 11, 12, 13, 14, 15, 16, 17] or predicting motor scores [18, 19, 20, 21] but often overlook region-specific dynamics that are essential for forecasting volume loss and defining individual disease trajectories. In this study, we primarily answer two questions: *(1) What is the annual average percentage of atrophy in PD-related brain regions? (2) Can an AI model predict brain atrophy for an individual patient (personalized care)?*

To address the first question, we analyzed 197 PD patients with their 424 longitudinal brain volumes, we calculated the annualized rate of atrophy (ARA) for PD-related brain regions. We discussed them in Section 3.1, 3.2 and 3.3. For the second question, we developed LSTM [22] regression model and classification models to capture personalized, longitudinal profiles of regional atrophy in 29 brain regions and its evolution over time (Section 3.4). In addition to structural MRI measures these models integrate 122 clinical and biomarker features including motor and cognitive scores, demographic variables, and cerebrospinal fluid analysis. The classification model facilitates prediction whether a patient will show a prespecified percentage of ARA decline by the next visit.

Our approach serves as a proof-of-concept demonstrating feasibility of generating clinically meaningful, region-specific predictions of neurodegeneration in PD. As these tools continue to develop, they are expected to provide a valuable reference for mechanistic hypothesis generation, anticipating disease progression, and planning more personalized care.

## 2 Methods

### 2.1 Dataset and Cohort Structure

We collected a longitudinal multi-modal dataset comprising 197 unique patients with a confirmed diagnosis of PD from the Parkinson’s Progression Markers Initiative (PPMI) [23]. Each patient contributed between two and four clinical visits, resulting in 424 total time points collected over a follow-up period of two to five years, as shown in Table 1. Patients with fewer than two qualified structural MRIs and 105 patients with missing clinical records were excluded from the study. No demographic or clinical differences were observed between these removed individuals and the training cohort. At each visit, patients underwent both high-resolution T1-weighted 3D brain MRI on 1.5T or 3T scanners along with comprehensive clinical assessments. The clinical data included 122 structured features such as age, sex, cognitive scores including the Montreal Cognitive Assessment (MoCA) [24], motor evaluations using the MDS-UPDRS [25], or cerebrospinal fluid (CSF).

**Table 1:** Baseline Demographics and Clinical Characteristics.

| Characteristic | Value |
| --- | --- |
| Age — years |  |
| Median (range) | 64.1 (36–84) |
| <65 — no. (%) | 109 (56.1%) |
| 65–74 — no. (%) | 68 (32.5%) |
| ≥75 — no. (%) | 20 (9.4%) |
| Sex — no. (%) |  |
| Male | 135 (68.6%) |
| Female | 65 (31.4%) |
| Body-mass index (BMI) |  |
| Median (range) | 25.35 (16.8–43.4) |
| <18.5 — no. (%) | 4 (2.1%) |
| 18.5–24.9 — no. (%) | 84 (43.6%) |
| 25–29.9 — no. (%) | 80 (38.9%) |
| ≥30 — no. (%) | 27 (14.4%) |
| Race/Ethnicity — no. (%) |  |
| White | 178 (90.3%) |
| Black | 6 (3.1%) |
| Asian | 4 (2.4%) |
| Other (includes multi-racial) | 8 (3.8%) |
| Unknown / Not reported | 1 (0.5%) |
| Hoehn & Yahr stage — no. (%) ($N = 424$)<sup>1</sup> | |
| Stage 0 | 0 (0.0) |
| Stage 1 | 93 (21.9%) |
| Stage 2 | 280 (66%) |
| Stages 3–5 | 15 (3.5%) |
| Unknown | 36 (8.5%) |
| MoCA score ($N = 424$)<sup>1</sup> | |
| Median (IQR) | 27.0 (IQR 26.0–29.0) |

<sup>1</sup>MoCA and Hoehn & Yahr stage use a total of 424 patient records.

### 2.2 3D MRI Feature Extraction

All T1-weighted scans were processed using FastSurfer [26] a deep learning-based model of the FreeSurfer [27] segmentation pipeline [28]. The preprocessing pipeline included motion correction, bias field adjustment, skull stripping, and cortical and subcortical segmentation [29]. Each scan yielded a whole-brain parcellation consisting of 94 volumetric regions based on the DKT atlas [30], including both subcortical (e.g., caudate, putamen, thalamus) and cortical labels. All MRI scans were processed on high-performance GPU computing infrastructure. Longitudinal plots of regional volumes across visits revealed consistent and progressive atrophy in key brain regions implicated in PD pathophysiology. The full 216 extracted features (94 brain volumes and 122 clinical features) are discussed in Supplementary. We did not employ a separate ICV regressor; our use of within-subject percent change was chosen to correct for individual variation in baseline volume.

### 2.3 Model Design and Training Procedure

We developed classification models, XGBoost [31], that integrate both clinical features and regional brain volumes to classify whether a patient will have a specified ARA of volume decline in a brain region by the next year visit. We also constructed an LSTM model to predict the exact percentage of brain atrophy on the next visit. For the LSTM model, we used a single-branch recurrent architecture that ingests time-stamped numeric features (including regional volumes and clinical variables) as a sequence. Inputs were zero-padded and passed through a Masking layer, followed by a two-layer LSTM stack (128 and 64 units) with dropout and recurrent dropout of 0.2 in each layer. A final Dropout (0.4) was applied before the output heads. The network produced a multi-output regression [32] head that predicts the annualized % (region atrophy percentage) for each region.

### 2.4 Evaluation Metrics and Model Interpretability

The model’s predictive performance was evaluated using standard regression metrics. Mean absolute error (MAE) [33], coefficient of determination (*R*^2^) [34] and standard deviation (SD) [35] are used to quantify the average magnitude of prediction error across brain regions. Root mean squared error was calculated to emphasize larger deviations from true values. These metrics were computed for each brain region and averaged to obtain an overall performance score. To examine the model’s clinical utility, we conducted exploratory binary classification analyses by applying thresholds to predicted atrophy values as each brain region has a different ARA atrophy rate. AUROC [36], Sensitivity (Recall), Specificity [37] and Testing Accuracy were then used to assess the classification model’s capability. To improve interpretability and trustworthiness of the model, we employed SHapley Additive exPlanations (SHAP) [38] to analyze feature importance and generate global explanation plots to summarize feature influences.

### 2.5 Statistical Analysis

As PPMI collects data from multiple clinical sites, we split the dataset by site into 80% for training and 20% for testing for model generalization. This ensures that test data from a given site is not used in the training set. Within the training data, we used 5-fold cross-validation with grouped and stratified splits by clinical site. Decision thresholds were tuned (automatically selected by model) on validation folds to reach high sensitivity for clinical relevance. We then compared regions using fold means and their 95% confidence intervals. To relate regions to clinical status, we computed Spearman rank correlations [39] between 94 brain region volumes and MoCA and MDS UPDRS3. As the number of participants with both multi time point MRI and long-term outcomes such as incident dementia or PD MCI conversion was modest, our study was not powered for stable subgroup analyses that contrasted predicted atrophy trajectories across outcome strata. We therefore focused on overall model performance for regional atrophy prediction in early PD and treated structure function associations as hypothesis generating. Finally, we clarify that the data splitting was performed at the patient level, ensuring no overlap of subjects between sets. The 5-fold CV was strictly nested within the training set, and the test set was never seen during training.

## 3 Results

### 3.1 Annualized Rate of Atrophy (ARA)

Annualized rate of atrophy (ARA) was analyzed for PD-related regions to understand the average value of regional brain atrophy. The ARA values also can be used to classify whether an individual progression will be above average or lower.

As shown in Fig. 1, 94 brain regions are analyzed, and 29 regions’ ARA shows remarkable atrophy. We confirmed that the 29 findings are statistically significant (*p* < 0.05), meaning less than 5% probability that the observed atrophy patterns are noise or random fluctuations.

**Figure 1:**
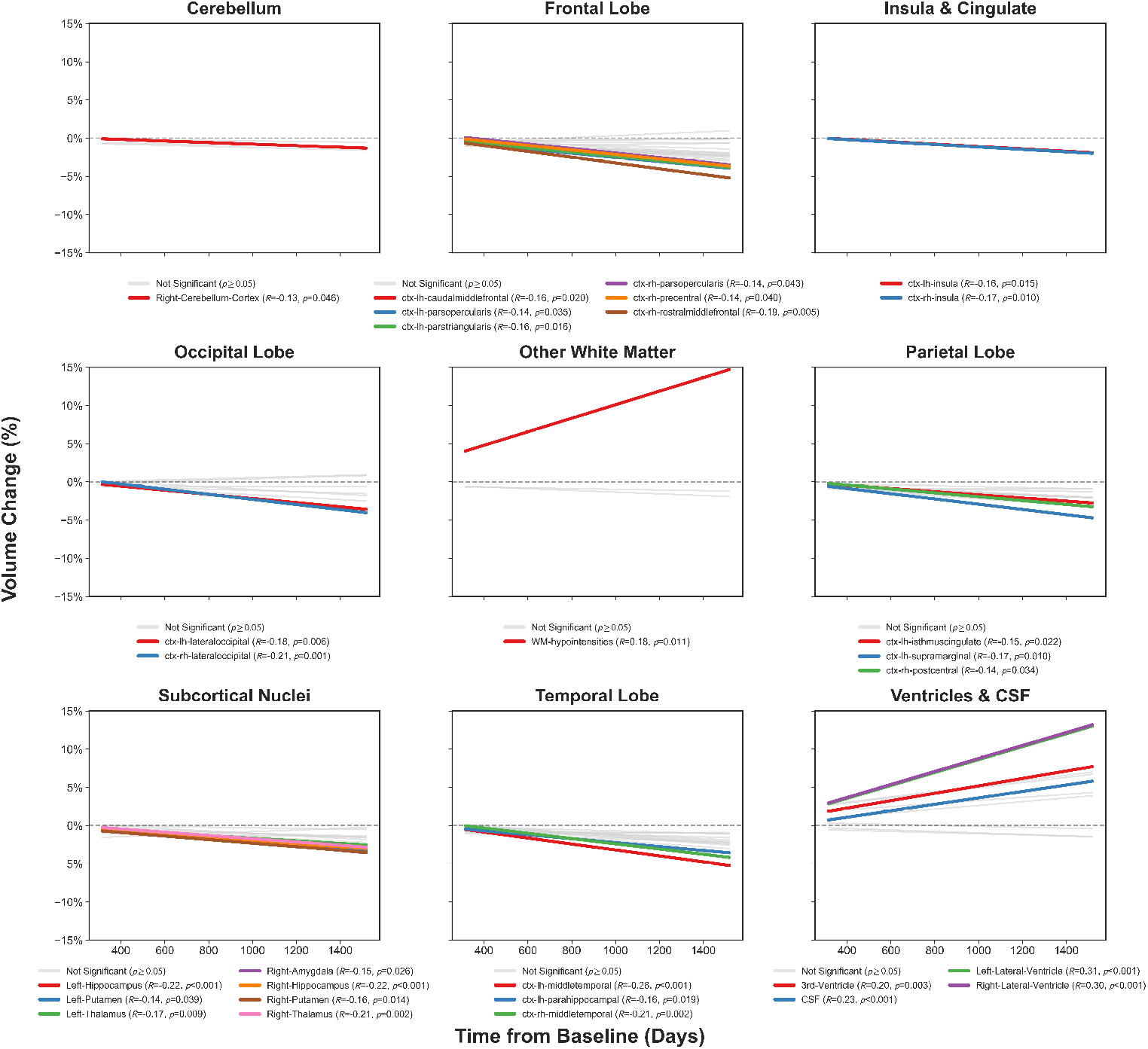
Annualized rate of atrophy in highlighted regions. Colored lines indicate significant volume changes (*p* < 0.05), while gray lines represent non-significant changes (*p* ≥ 0.05). Most regions show progressive volume loss, except for Ventricles & CSF, which show volume expansion.

We observed that fluid expansion markers showed the most significant longitudinal changes, making them robust indicators of global atrophy. Notably, white matter hypointensities (WMH) had the highest ARA at +3.23%/year. The ventricular system, with the right and left lateral ventricles expanding at + 3.10%/year and + 3.09%/year, respectively. The third ventricle (+1.77%/year) and overall CSF volume (+1.54%/year) also demonstrated significant yearly increases. Frontal lobe regions were also among the most rapidly degenerating, especially those linked to motor control and the rates also consistent with the latest findings. The right rostral middle frontal gyrus shows an ARA of −1.38%. We found that both the right precentral gyrus (−1.07%/year) and the left caudal middle frontal gyrus (−1.02%/year) exceeded the 1% ARA. Language-related frontal regions were also notably affected, including the right pars opercularis (−1.08%/year), left pars triangularis (−1.01%/year), and left pars opercularis (−0.99%/year). Additionally, the right rostral anterior cingulate cortex declined by −1.09%/year. All the twenty-nine region ARA values are shown in Table 2.

**Table 2:** Model performance for all PD-related regions.

| Region | ARA(%) | AUC (95% CI) | P-Val | ACC | SENS | PREC | LSTM MAE | LSTM $R^2$ | LSTM SD |
| --- | --- | --- | --- | --- | --- | --- | --- | --- | --- |
| ctx-rh-precentral | -1.07 | 0.842 [0.832-0.852] | <.0001 | 0.884 | 0.937 | 0.823 | 0.537 | 0.533 | 0.927 |
| Left- Lateral-Ventricle | 3.09 | 0.905 [0.897-0.912] | <.0001 | 0.871 | 0.87 | 0.875 | 0.349 | 0.797 | 0.548 |
| Right- Lateral-Ventricle | 3.1 | 0.914 [0.906-0.922] | <.0001 | 0.843 | 0.871 | 0.848 | 0.395 | 0.789 | 0.533 |
| ctx-rh-insula | -0.59 | 0.834 [0.822-0.846] | <.0001 | 0.795 | 0.834 | 0.726 | 0.502 | 0.591 | 0.874 |
| ctx-lh-middletemporal | -1.43 | 0.803 [0.789-0.817] | <.0001 | 0.815 | 0.809 | 0.751 | 0.443 | 0.586 | 0.730 |
| 3rd-Ventricle | 1.77 | 0.826 [0.816-0.837] | <.0001 | 0.81 | 0.789 | 0.852 | 0.370 | 0.809 | 0.432 |
| Right-Cerebellum-Cortex | -0.37 | 0.822 [0.812-0.833] | <.0001 | 0.818 | 0.801 | 0.835 | 0.354 | 0.802 | 0.431 |
| ctx-rh-postcentral | -0.93 | 0.842 [0.832-0.852] | <.0001 | 0.796 | 0.77 | 0.806 | 0.549 | 0.507 | 0.838 |
| ctx-lh-caudalmiddlefrontal | -1.02 | 0.802 [0.790-0.813] | <.0001 | 0.791 | 0.765 | 0.807 | 0.532 | 0.590 | 0.759 |
| Left- Putamen | -0.76 | 0.787 [0.775-0.800] | <.0001 | 0.786 | 0.794 | 0.800 | 0.529 | 0.508 | 0.717 |
| ctx-lh-supramarginal | -1.25 | 0.752 [0.738-0.765] | <.0001 | 0.808 | 0.784 | 0.826 | 0.504 | 0.459 | 0.687 |
| ctx-rh-rostralmiddlefrontal | -1.38 | 0.825 [0.813-0.837] | <.0001 | 0.774 | 0.83 | 0.702 | 0.545 | 0.547 | 0.855 |
| ctx-lh-insula | -0.57 | 0.783 [0.769-0.798] | <.0001 | 0.798 | 0.751 | 0.759 | 0.480 | 0.609 | 0.789 |
| Right- Putamen | -0.85 | 0.779 [0.768-0.790] | <.0001 | 0.781 | 0.764 | 0.784 | 0.555 | 0.481 | 0.947 |
| ctx-lh-parsopercularis | -0.99 | 0.815 [0.804-0.826] | <.0001 | 0.759 | 0.804 | 0.783 | 0.505 | 0.387 | 0.808 |
| Right- Hippocampus | -0.71 | 0.819 [0.807-0.831] | <.0001 | 0.803 | 0.823 | 0.817 | 0.439 | 0.610 | 0.818 |
| Left- Thalamus | -0.62 | 0.760 [0.745-0.775] | <.0001 | 0.767 | 0.64 | 0.847 | 0.382 | 0.777 | 0.530 |
| Right- Thalamus | -0.78 | 0.764 [0.750-0.779] | <.0001 | 0.776 | 0.653 | 0.770 | 0.455 | 0.678 | 0.658 |
| Left- Hippocampus | -0.81 | 0.795 [0.784-0.807] | <.0001 | 0.77 | 0.806 | 0.731 | 0.353 | 0.702 | 0.592 |
| ctx-rh-lateraloccipital | -1.22 | 0.760 [0.749-0.772] | <.0001 | 0.767 | 0.774 | 0.684 | 0.427 | 0.707 | 0.625 |
| ctx-lh-lateraloccipital | -0.99 | 0.779 [0.766-0.791] | <.0001 | 0.758 | 0.729 | 0.744 | 0.417 | 0.692 | 0.623 |
| ctx-rh-parsopercularis | -1.08 | 0.744 [0.732-0.756] | <.0001 | 0.721 | 0.778 | 0.663 | 0.539 | 0.515 | 0.839 |
| ctx-rh-middletemporal | -1.25 | 0.748 [0.736-0.760] | <.0001 | 0.801 | 0.665 | 0.683 | 0.476 | 0.633 | 0.756 |
| ctx-lh-isthmuscingulate | -0.75 | 0.711 [0.698-0.723] | <.0001 | 0.708 | 0.729 | 0.682 | 0.492 | 0.502 | 0.677 |
| ctx-lh-parstriangularis | -1.01 | 0.738 [0.727-0.750] | <.0001 | 0.705 | 0.814 | 0.653 | 0.577 | 0.578 | 0.757 |
| ctx-lh-parahippocampal | -0.94 | 0.685 [0.669-0.701] | <.0001 | 0.736 | 0.689 | 0.741 | 0.506 | 0.467 | 0.712 |
| CSF | 1.54 | 0.641 [0.624-0.657] | <.0001 | 0.698 | 0.684 | 0.658 | 0.544 | 0.422 | 0.718 |
| WM-hypointensities | 3.23 | 0.618 [0.606-0.631] | <.0001 | 0.653 | 0.578 | 0.790 | 0.279 | 0.800 | 0.408 |
| Right- Amygdala | -0.95 | 0.615 [0.599-0.630] | <.0001 | 0.602 | 0.708 | 0.531 | 0.531 | 0.520 | 0.906 |

**Table 3:**
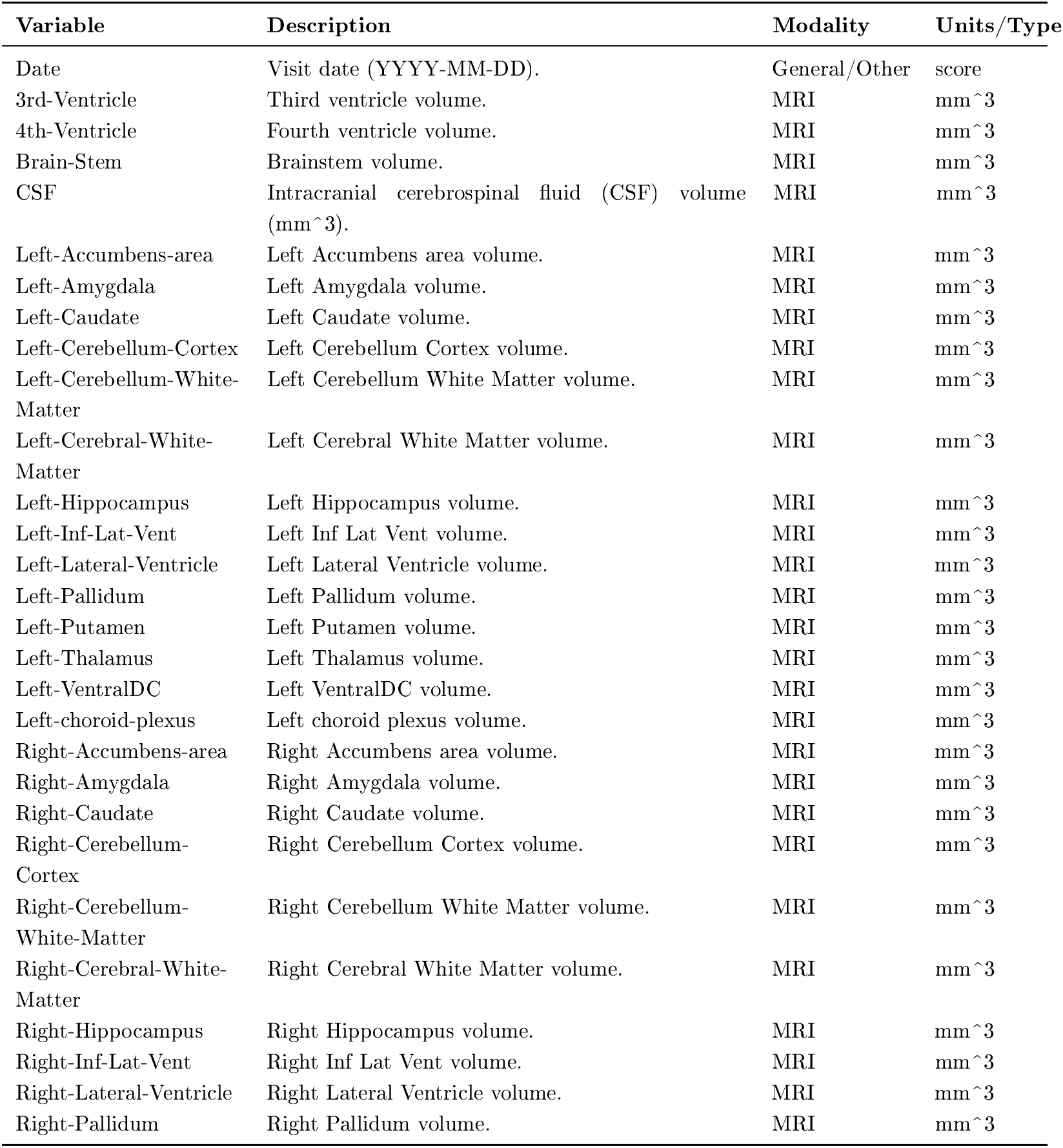
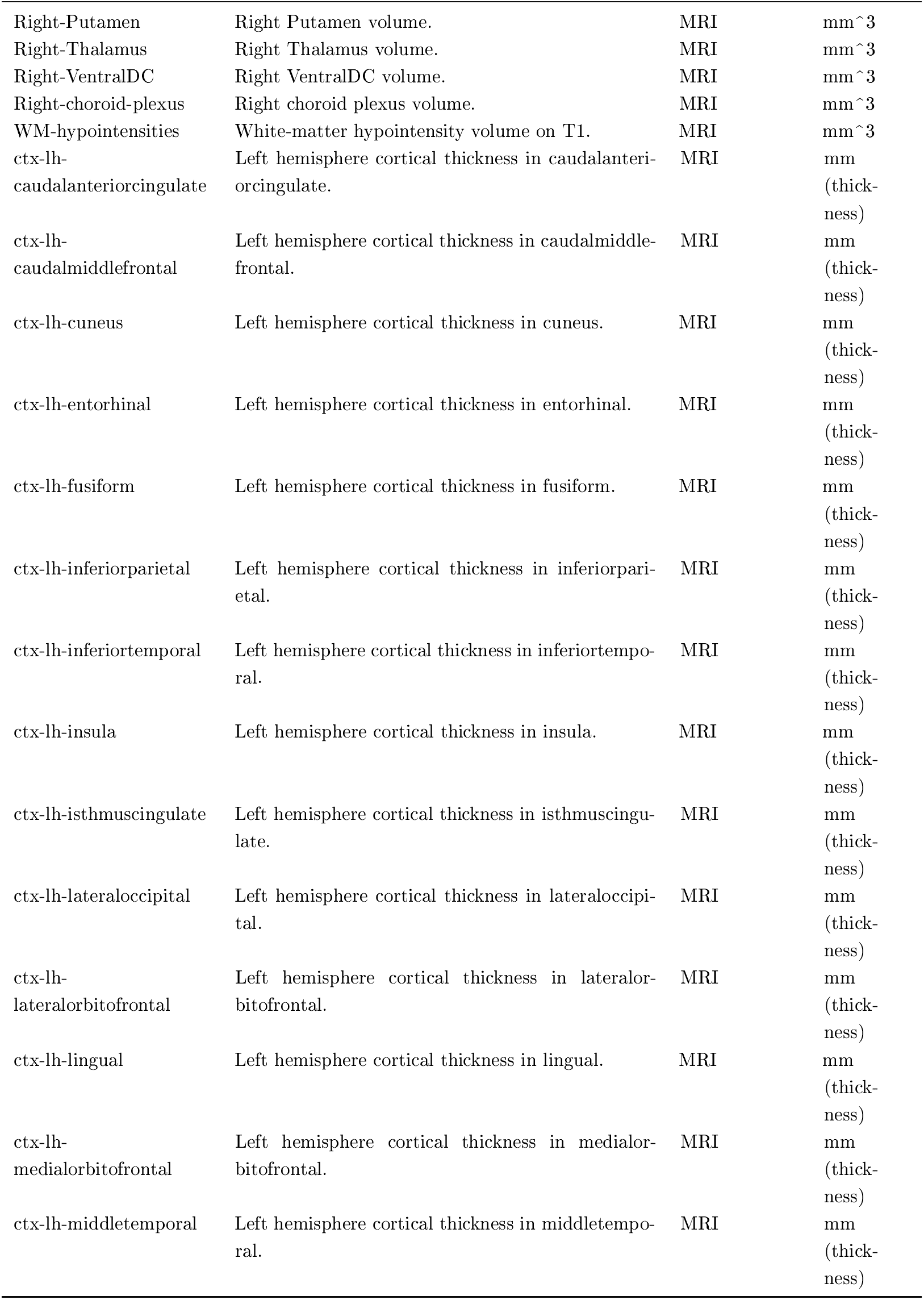
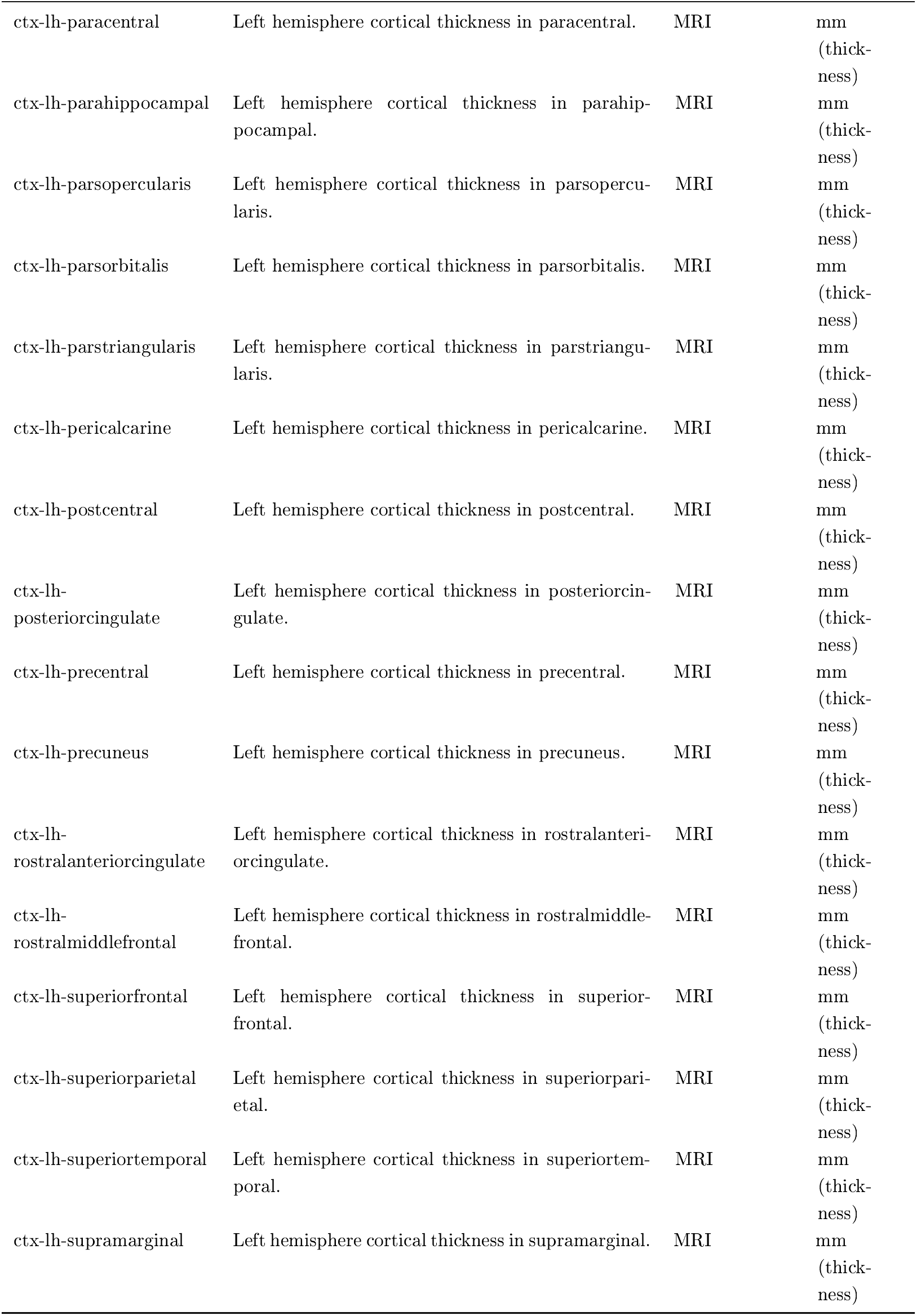
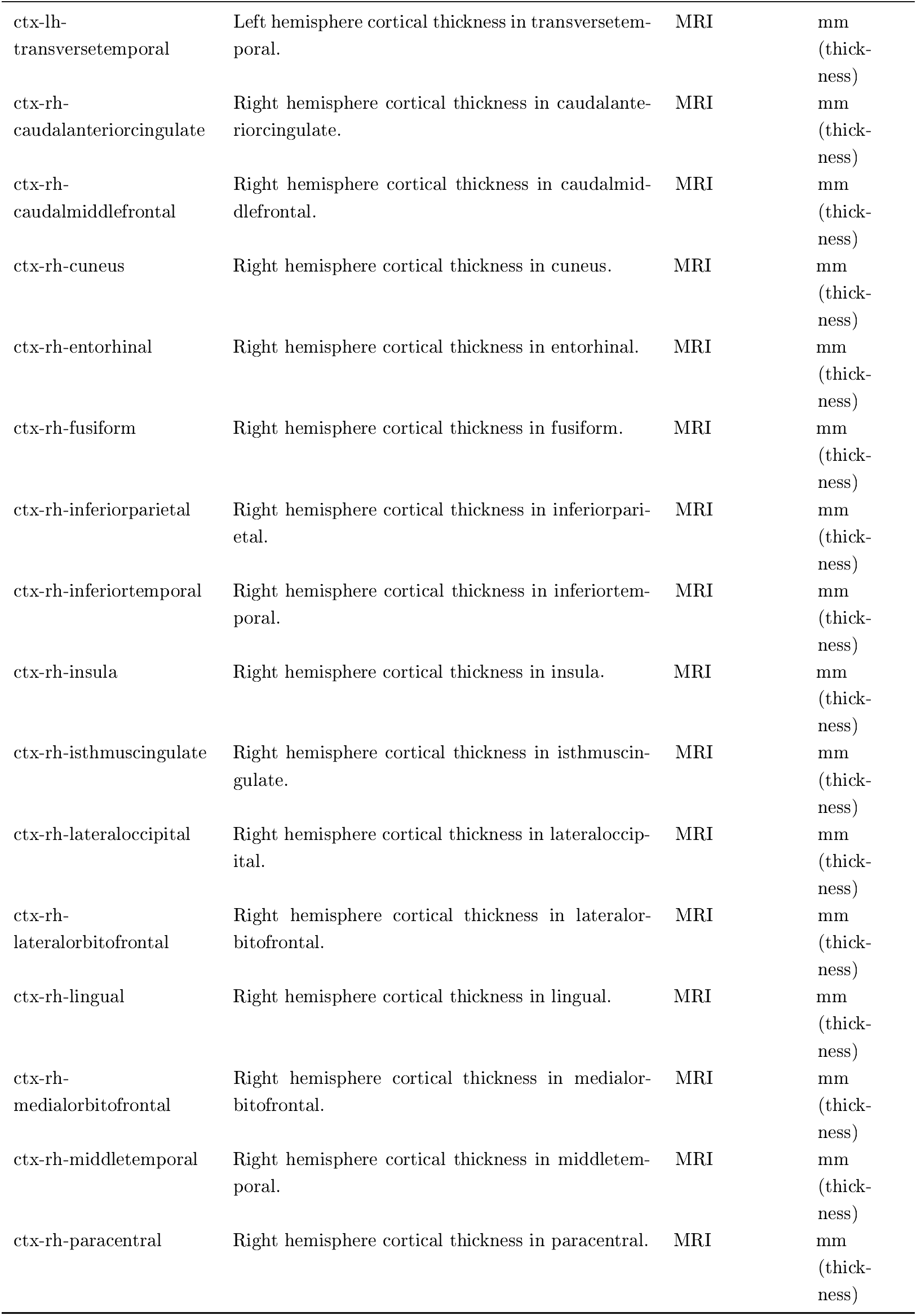
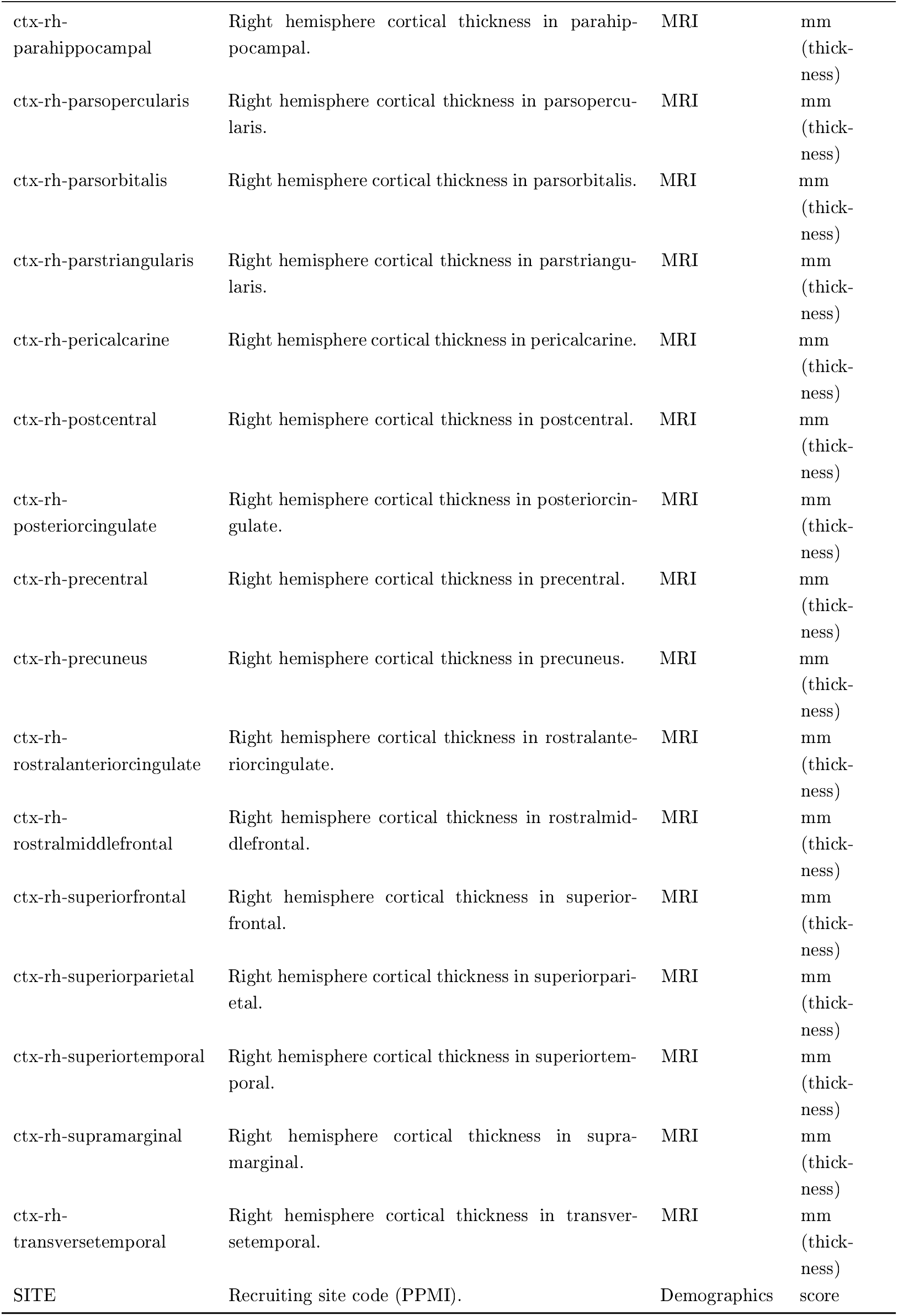
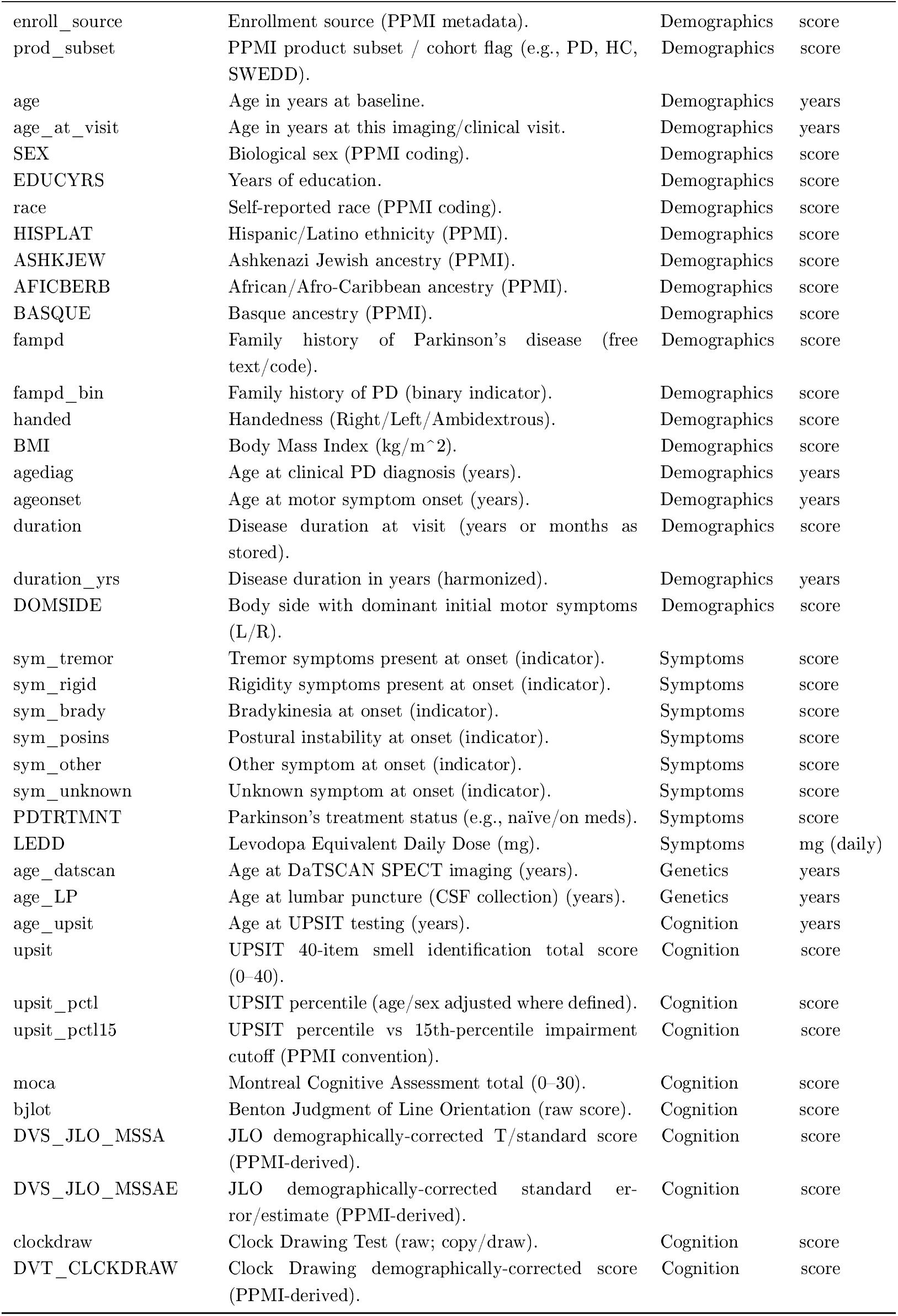
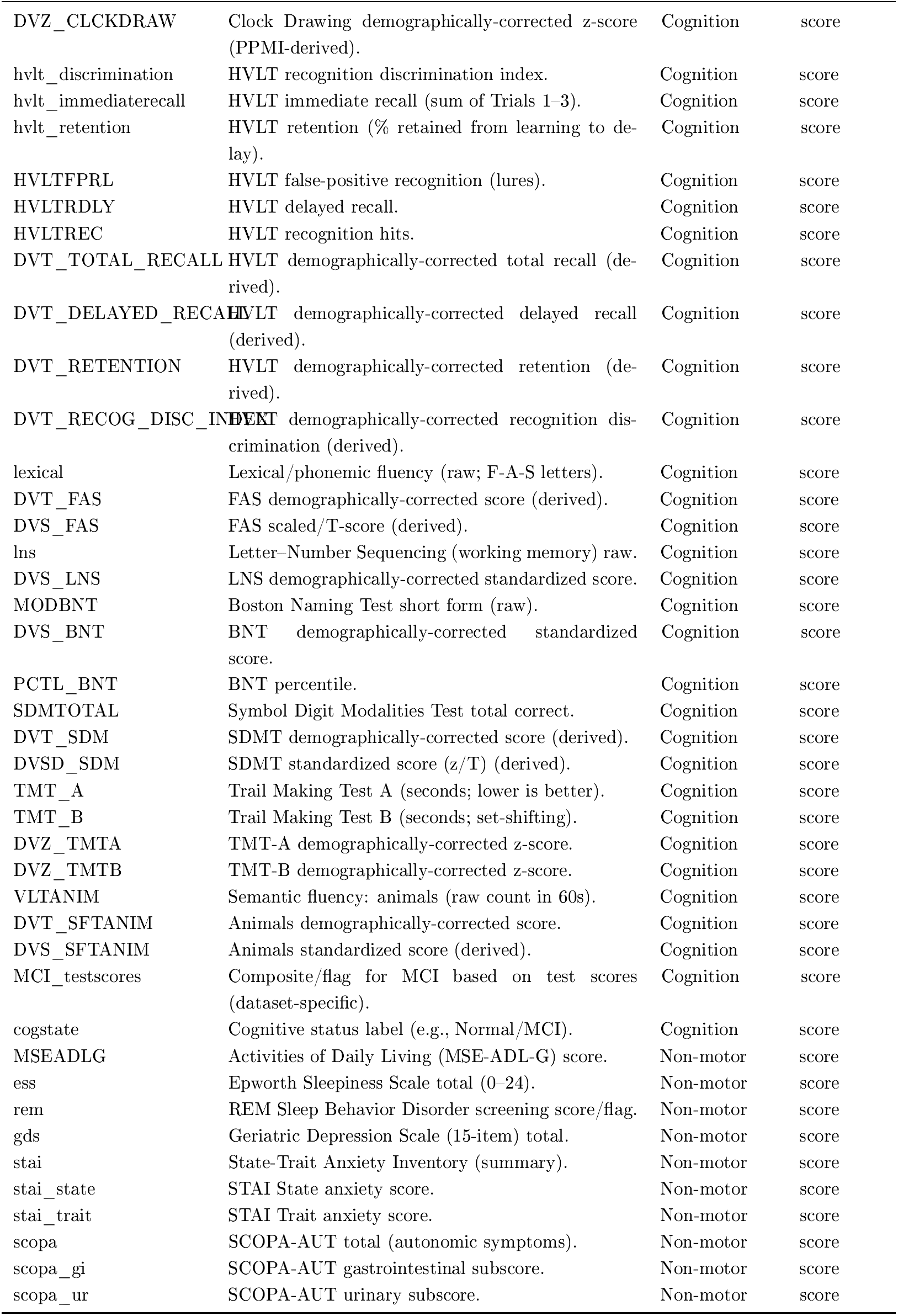
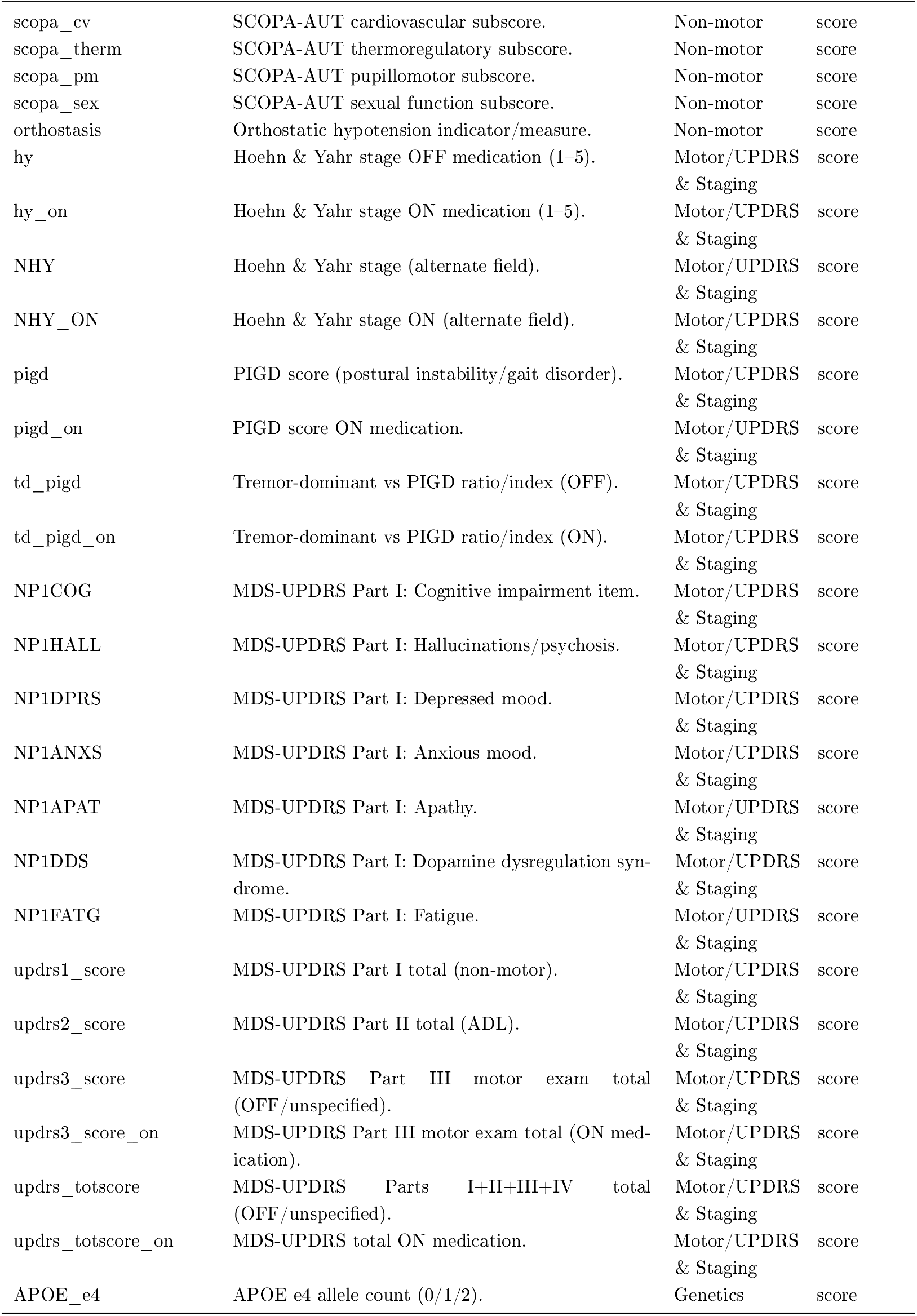
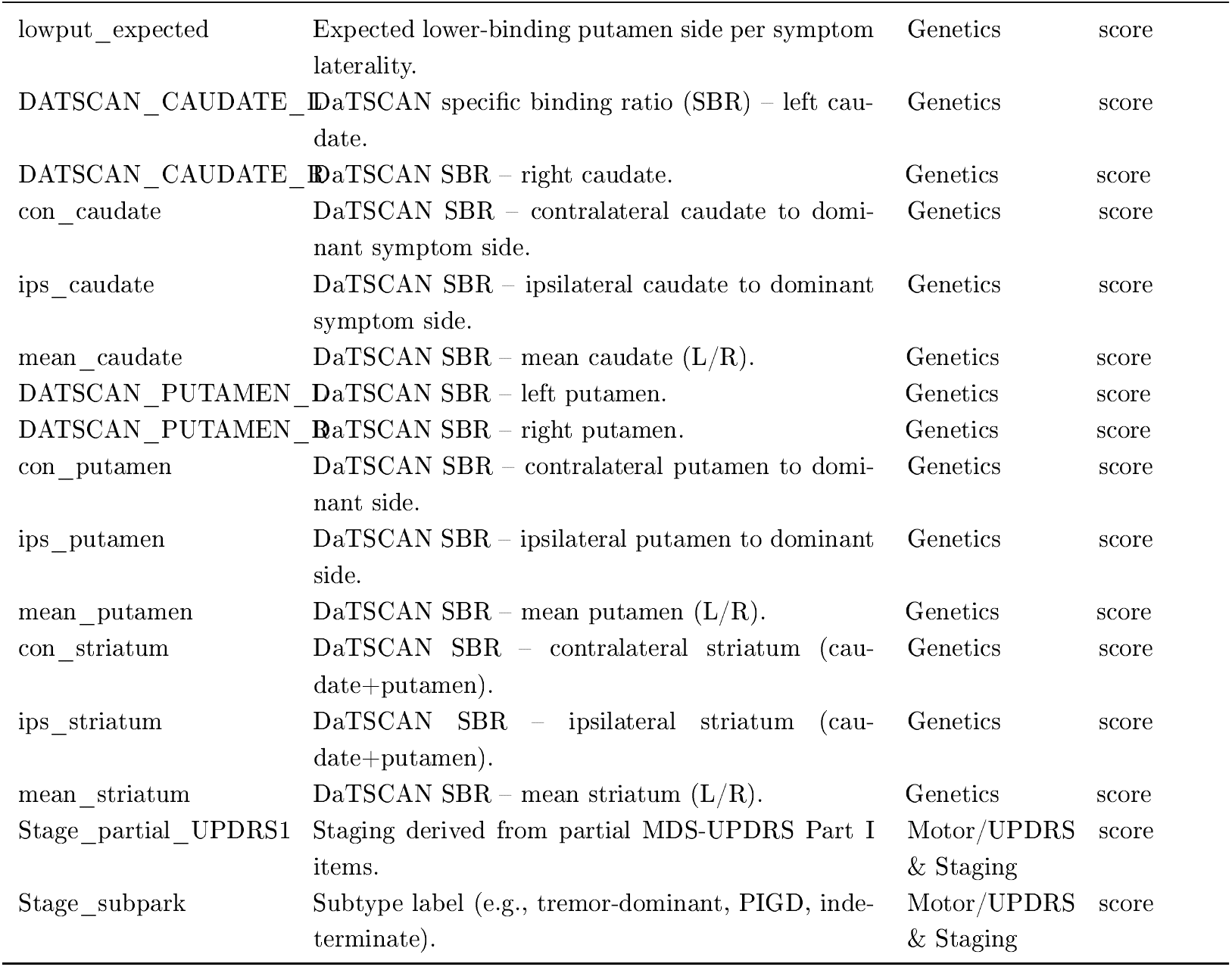
Feature Dictionary for Imaging, Clinical, and Neuropsychological Variables.

### 3.2 Distribution of Percentage Change in Brain Regions

In Fig. 2, we have shown the distributions of percent volume change from baseline aggregated across all visits in 197 participants as a violin plot. The mean of most regions exhibits a small negative central tendency, consistent with mild atrophy. Striatal and limbic structures including left and right putamen, caudate, amygdala, and hippocampus have median changes of approximately −1% to −5%, with left-skewed tails extending to about −30%. Ventricular regions display positive medians and broad right-skewed spreads, indicating significant percentage enlargement in a subset of participants, with maximum enlargement over time of around 35%. The brainstem centers on a slight loss with relatively modest dispersion.

**Figure 2:**
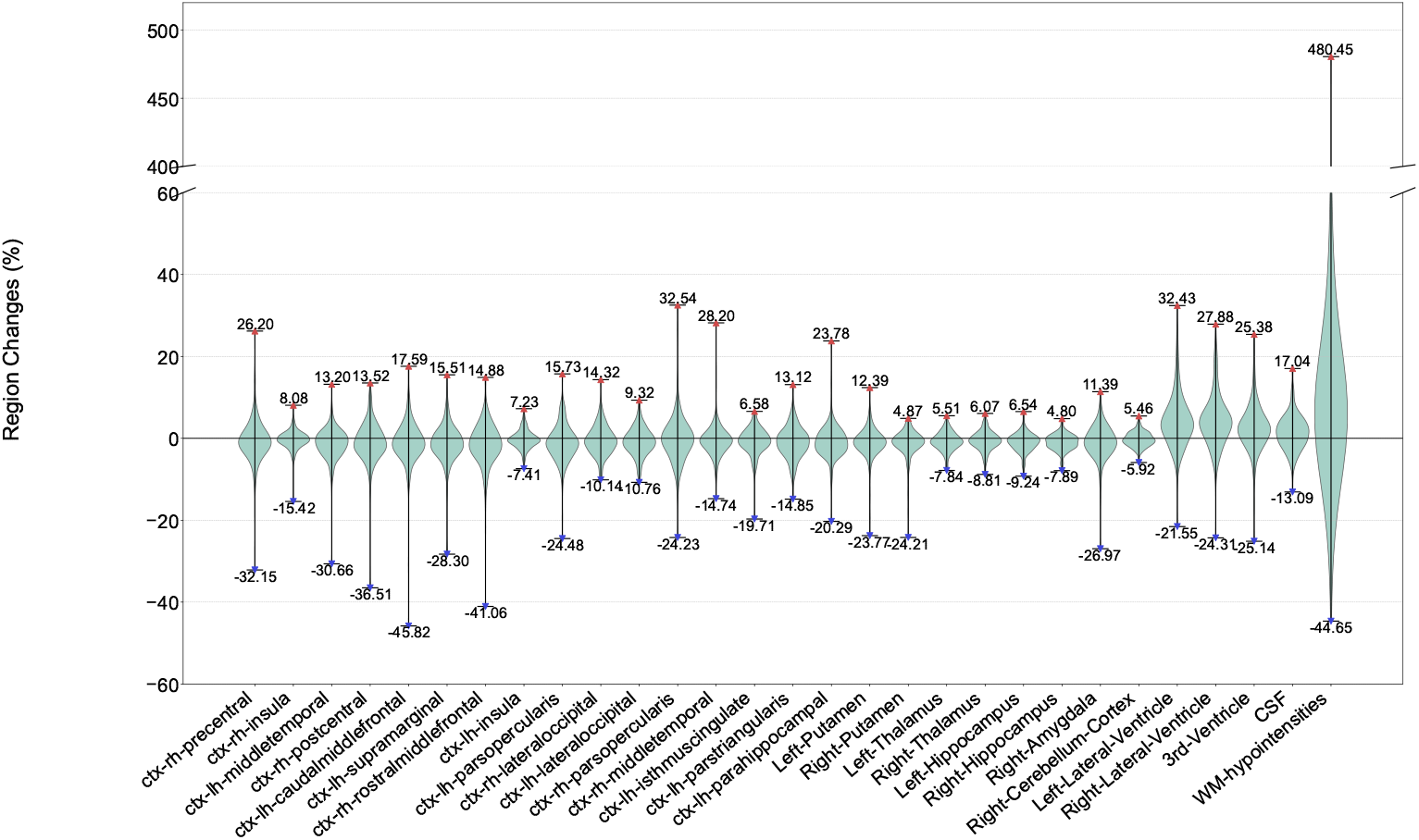
Percentage atrophy distribution of regional brain volumes. Violin plots illustrate the distribution of volume changes (%) across specific brain regions. Red and blue markers represent the maximum and minimum percentage changes for each region, respectively.

We found the white matter (WM) hypointensities reached a maximum value of 480.45%, indicating a high heterogeneity of neurodegenerative patterns within the parkinsonian brain. The value is caused by the small baseline in healthy controls, for example, a patient in our cohort shows a baseline volume is 1834 mm^3^ increased to 6679 mm^3^ after a year.

### 3.3 Correlation Between Brain Region Atrophy and MDS-UPDRS3/MoCA

We computed Spearman correlations between volumetric change across 94 brain regions and two key clinical measures (MoCA and MDS-UPDRS3). The analysis identified 52 regions showing positive or negative correlations (Fig. 3), with 13 regions overlapping with these two clinical measures (e.g., CSF, WM hypointensities, ventricle, thalamus, or amygdala).

**Figure 3:**
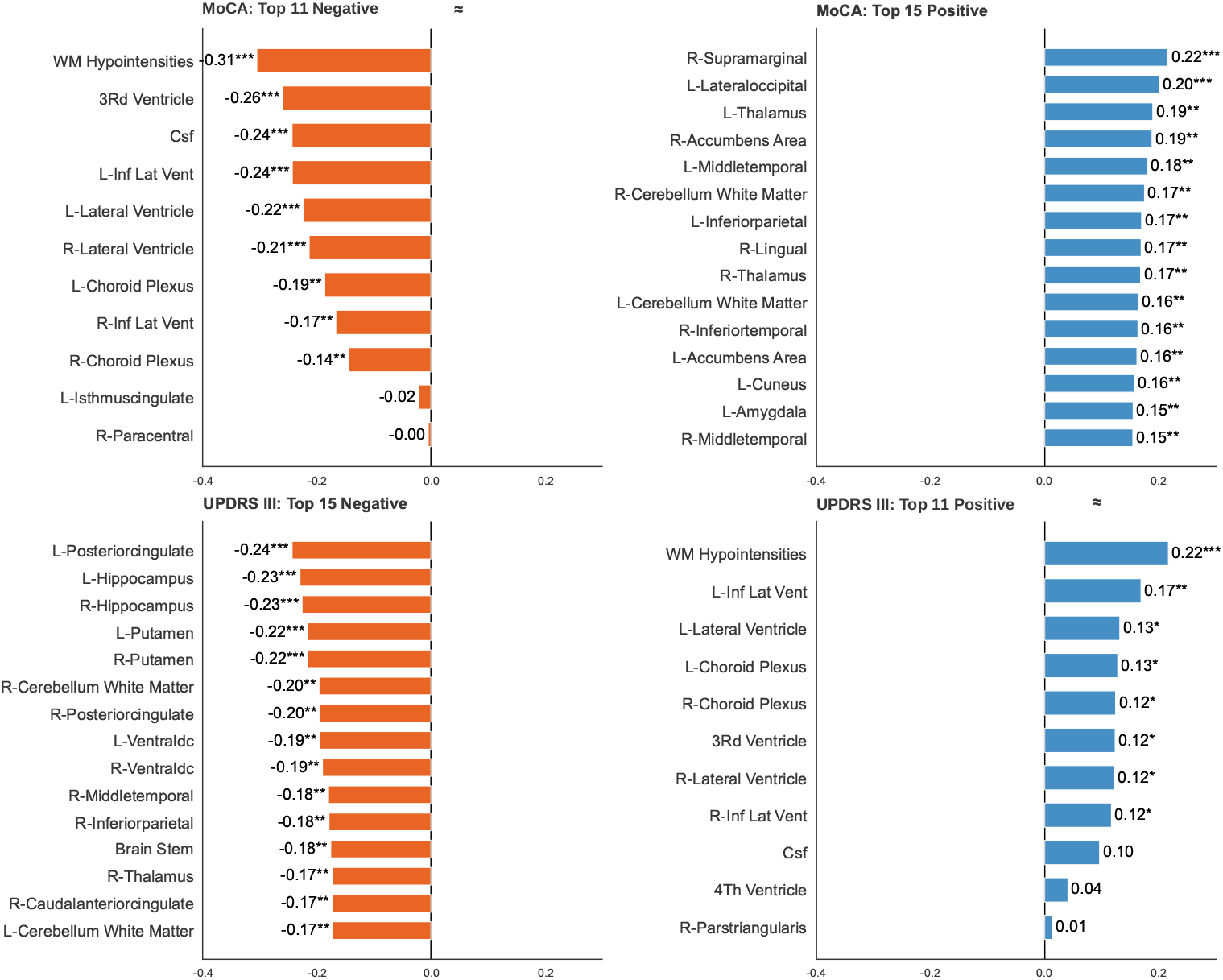
Correlation between brain region atrophy and MDS-UPDRS3/MoCA. Bar charts show the top regions correlated with MoCA and MDS-UPDRS III scores. Orange and blue bars represent negative and positive correlations, respectively. Asterisks denote statistical significance: \**p* < 0.05, * \**p* < 0.01, * * \**p* < 0.001.

Fig. 3 reports Spearman correlations between regional brain volumes and clinical scores in PD. For the MoCA, the largest magnitude associations are negative and involve cerebrospinal fluid spaces. Greater 3rd-ventricular, lateral ventricular, and CSF volumes correlate with lower MoCA scores, consistent with poorer cognition when ventricular and CSF spaces are enlarged. Positive correlations with MoCA are largely confined to cortical territories and are small, suggesting that larger regional volumes will have slightly better cognitive performance, but the effects are modest. For the MDS-UPDRS3, the pattern generally reflects motor worsening correlating with volume loss. Larger ventricular and CSF volumes show the strongest positive correlations with higher MDS-UPDRS3 scores, indicating worse motor scores when fluid spaces are expanded. The top negative correlations include hippocampal and other cortical or subcortical regions, meaning that larger volumes are associated with better motor performance and, conversely, that smaller volumes accompany greater motor impairment.

### 3.4 Multi-Modal Data Enables High Accuracy

#### Classification Model

We trained deep learning models with grouped, patient-stratified 5-fold cross-validation, where a decision threshold was applied to fit regions with higher sensitivity. As shown in Table 2, the AUROC of the Left and Right Lateral Ventricle reached 0.905 and 0.914, respectively, indicating they are the best markers for classification. The sensorimotor-related regions, including the precentral and postcentral gyri, showed strong performance with AUROC values above 0.80. Cognitive and executive-related regions, such as the rostral and caudal middle frontal cortices as well as the insula, also achieved AUROC values exceeding 0.80. In addition, language-related regions in the left hemisphere, including the middle temporal gyrus and the pars opercularis, demonstrated similarly high classification performance. Typically, if the model predicts that motor cortical regions exceed the ARA value, it indicates greater disease severity and progression in PD. Finally, all the models’ AUROC p-value are < 0.0001, which indicates our models learned real patterns from the dataset.

#### Regression Model

The LSTM model shows strong predictive power for brain regions related to Parkinson’s disease. Most regions have an *R*^2^ score between 0.5 and 0.8. This suggests that the model effectively captures the model features. As shown in Table 2, our model also performs best on the ventricular system of the brain. For example, the 3rd-Ventricle and Left-Lateral-Ventricle both have *R*^2^ scores near 0.8. These areas usually change steadily over time, so the model tracks their growth easily. The MAE for these regions is also low, which proves the predictions are accurate.

Predicting cortical regions like the insula and temporal lobe is more diffcult. These areas show lower *R*^2^ scores and higher error rates. The CSF and the parsopercularis have *R*^2^ values below 0.43, suggesting more noise in that data. The SD values also highlight some instability in the model. A high SD in the Right Putamen and Right Amygdala mean the model’s performance varies a lot between different subjects. Overall, the LSTM model is reliable for deep structures but needs more work for complex surface regions.

### 3.5 Feature Analysis and Model Explanation

Fig. 4a illustrates how different brain features contribute to the 29 regional models. We identified the most consistent predictors by averaging the absolute SHAP values across the entire model set. Every feature in this top 20 list is prefixed with *“*Diff’ to indicate that the model is primarily driven by longitudinal changes over time instead of static baseline measurements. The results clearly show that the lateral ventricles are the most influential predictors in this study. The high importance of both the left and right ventricles suggests that their expansion over time is a more robust indicator for the model than localized changes in the cortex. This is followed by the left cerebellum cortex and the right precentral gyrus, which highlights the significant role of motor-related and cerebellar pathways in the classification process. Several subcortical nodes also appear prominently in the top ten rankings. Specifically, the model relies heavily on changes in the hippocampus, thalamus, and putamen. The consistent ranking of these regions across all 29 models reinforces their status as core biomarkers for the neurobiological progression under investigation. While cortical areas such as the superior temporal gyrus and precuneus remain relevant, their contribution is secondary to the more pronounced signals observed in the ventricles and deep brain structures.

**Figure 4:**
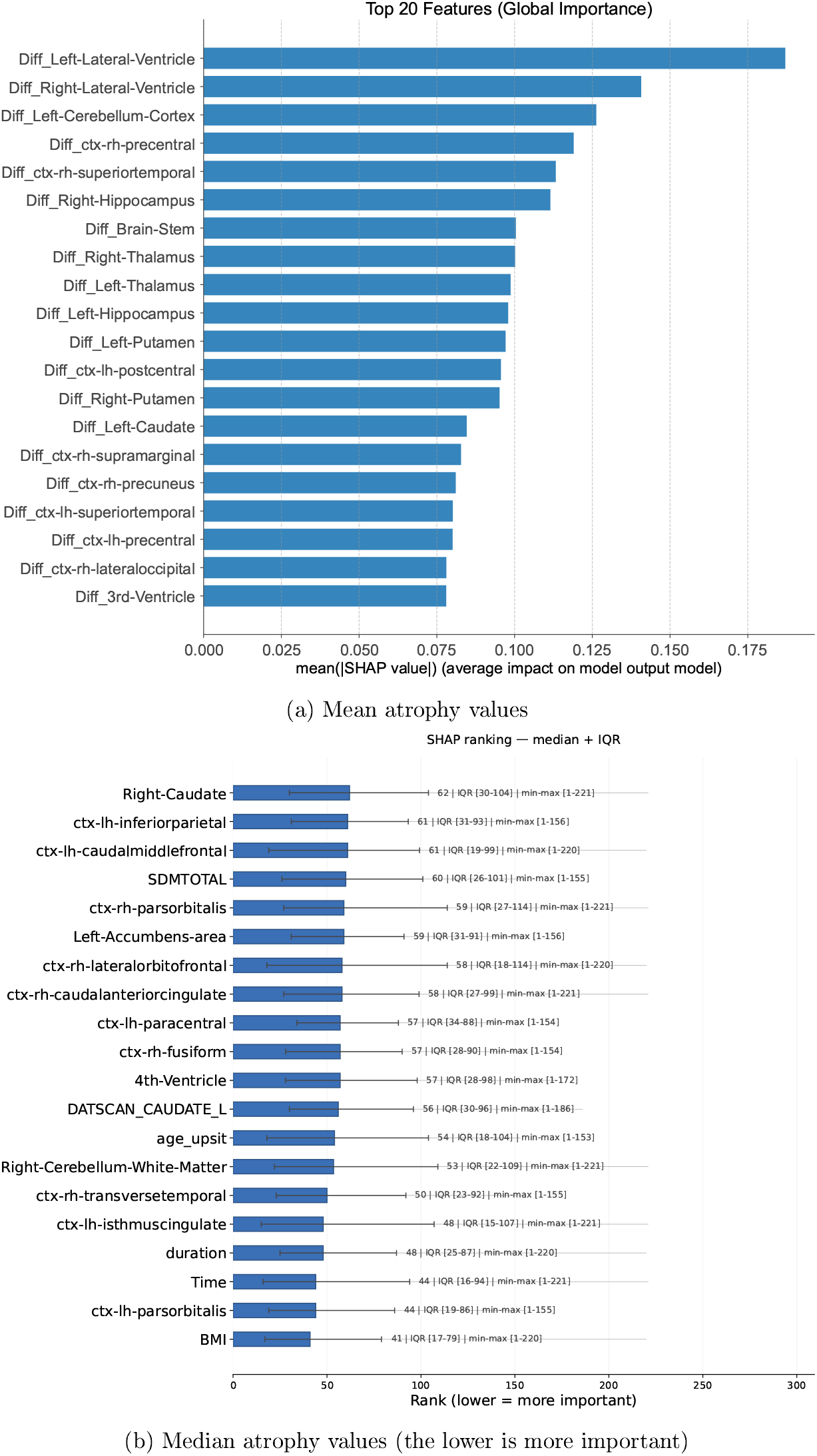
Global feature importance and ranking across models. (a) Top 20 features ranked by their mean absolute SHAP values, representing the average impact on model output. (b) Median ranking and interquartile range (IQR) of features across 29 models. Lower rank values indicate higher importance.

We also selected median value across 29 models to rank the model feature importance, as shown in Fig. 4b. Our method is designed to prevent outliers from the dataset. First, we collect 29 rank values for one feature, such as the BMI. We get one ranking array from each of the 29 models of BMI. Next, we sort them in ascending order, from the most important rank (lowest number) to the least important rank (highest number). Since we have 29 models, meaning that the median value is 15th. The result shows that the BMI median value is 41. Therefore, the 41 for BMI signifies that at least half (15 or more) of the 29 trained models assigned BMI a rank of 41 or lower (more important).

The ranking demonstrates that some clinical features, such as BMI, time, duration days, and upsit, play an imporant role in model training. These clinical importance of features surpassed many structural brain metrics, such as, ventricular volume, indicating their importance in predicting neurodegeneration.

## 4 Discussion and Limitations

### Significance

The present study demonstrates, for the first time, that region-specific trajectories of brain atrophy in Parkinson’s disease can be accurately forecast using multimodal data. Across 29 brain regions, our models achieved high predictive performance, particularly in the different ventricle, basal ganglia, and hippocampal which are most closely linked to PD pathology. These forecasts not only reproduce known patterns of degeneration, but quantify them prospectively, thereby identifying which regions are most likely to decline for a given patient. By combining longitudinal MRI, clinical, and biomarker data, the model successfully integrates disease heterogeneity into individualized and anatomically grounded predictions.

### Limitations

Brain segmentation errors remain a technical constraint. Accurate segmentation of small brain regions remains a limiting factor in volumetric modeling. Because minor boundary errors can disproportionately distort estimated volumes, deep learning-based solutions require exceptionally high precision in these structures. For example, the hippocampus and amygdala, which are among the smallest structures in our analysis, showed suboptimal segmentation performance.

Basal ganglia emphasis emerges de novo. Across all the models, feature importance concentrated in canonical sensorimotor and limbic-striatal circuits. Cortical contributors included left postcentral gyrus, bilateral insula, and bilateral precentral and postcentral cortices, alongside subcortical nodes such as putamen and accumbens. Brainstem and cerebellar cortex also showed substantial importance, consistent with motor and cerebellar-brainstem involvement in PD. Ventricular and choroid plexus measures ranked in the middle to lower range, suggesting that diffuse atrophy and cerebrospinal fluid related changes were less informative for group discrimination than regionally specific tissue measures. Amygdala and superior temporal cortices ranked even lower. Taken together, the importance pattern prioritizes sensorimotor and striatal pathways, consistent with established PD neurobiology, and supports the face validity of the classifiers. Crucially, the apparent basal ganglia prominence emerged de novo: no region-level priors or hand-crafted weights were imposed.

Our sample has inherent limitations. A central motivation for this work is to move toward individual level prognosis. In the present PPMI cohort, however, the subset of participants with both serial T1 imaging and well characterized downstream outcomes such as incident dementia or PD MCI conversion remains relatively small once stratified, which limits our ability to estimate stable group specific trajectories or to define volumetric cut points that correspond to clinically meaningful changes in MoCA or MDS-UPDRS3. Rather than present unstable subgroup curves, we chose to focus this proof-of-concept study on accurate prediction of regional atrophy across the cohort and on mapping predicted atrophy to cross sectional clinical measures. The PPMI cohort also predominantly consists of patients with early PD enrolled at academic movement disorder centers, who are largely Caucasian and highly educated. These findings may not generalize to older or more comorbid patients, those treated in community settings, or more ethnically and socioeconomically diverse PD populations. For these reasons, linking these volumetric forecasts to individual patient outcomes, such as cognitive decline or motor disability, will require larger and more heterogeneous datasets. It will be a key focus of our ongoing work in both internal institutional cohorts and external collaborations.

Clinical interpretation remains central to our analysis. In PD the MDS-UPDRS3 and MoCA scores are well-established measures of motor and cognitive function, respectively. Interestingly, our model assigned minimal weights to these clinical features when predicting regional brain volumes. It is important to note that this does not mean that brain structure is unrelated to functional decline. When we examined structure to function relationships outside the model, strong correlations reemerged between basal ganglia and limbic atrophy and worsening MDS-UPDRS3 and MoCA scores (Fig. 3), in line with known neuroanatomic-clinical relationships. Median atrophy ranking also shows the importance ranking of clinical features (Fig. 4b). The apparent disconnect reflects a key distinction: within-model feature importance measures contribution to a specific prediction target, whereas biological relevance must be validated through independent structure-function analyses. In this context, regional atrophy remains a meaningful substrate of PD progression, even if its direct contribution to volume prediction was limited. This underscores the need for complementary approaches that integrate model-derived metrics, statistical validation, and clinical expertise to understand PD mechanisms. We also note that structure-function correlations remain observational rather than demonstrating direct causal links.

## 5 Conclusions

This work establishes an initial proof of concept for modeling longitudinal, region-specific neurodegeneration in PD. Region-specific prediction of atrophy offers not only a personalized estimate of disease progression but also helps define the network architecture of degeneration in PD. These forecasts could identify both fast and slow progressors early in the disease course, when disease-modifying therapies could have the most long-term impact. Stratifying analyses by expected structural change within different circuits will allow hypothesis testing regarding why outcomes, motor and nonmotor, differ and diverge among patients. At present the primary value of these models is mechanistic and hypothesis generating rather than providing actionable prognostic guidance for individual patients. Over time, the true power of these models will emerge through integration with other modalities (diffusion imaging, connectivity mapping, and genetics) to create a unified multimodal understanding of PD progression.

## Data Availability

All data produced in the present study are available upon reasonable request to the authors

## Supplementary

